# The far side of the COVID-19 epidemic curve: local re-openings and re-closings based on globally coordinated triggers may work best

**DOI:** 10.1101/2020.05.10.20097485

**Authors:** Vadim A. Karatayev, Madhur Anand, Chris T. Bauch

**Affiliations:** School of Environmental Sciences, University of Guelph, Guelph, Ontario, Canada; Department of Applied Mathematics, University of Waterloo, Waterloo, Ontario, Canada

## Abstract

In the late stages of an epidemic, infections are often sporadic and geographically distributed. Spatially structured stochastic models can capture these important features of disease dynamics, thereby allowing a broader exploration of interventions. Here we develop a stochastic model of severe acute respiratory syndrome coronavirus 2 (SARS-CoV-2) transmission amongst an interconnected group of population centres representing counties, municipalities and districts (collectively, “counties”). The model is parameterized with demographic, epidemiological, testing, and travel data from Ontario, Canada. We explore the effects of different control strategies after the epidemic curve has been flattened. We compare a local strategy of re-opening (and re-closing, as needed) schools and workplaces county-by-county according to triggers for county-specific infection prevalence, to a global strategy of province-wide re-opening and re-closing according to triggers for province-wide infection prevalence. We find that the local strategy results in a similar number coronavirus disease (COVID-19) cases but significantly fewer person-days of closure, even under high inter-county travel scenarios. However, both cases and person-days lost to closure rise when county triggers are not coordinated and when testing rates vary among counties. Finally, we show that local strategies can also do better in the early epidemic stage but only if testing rates are high and the trigger prevalence is low. Our results suggest that pandemic planning for the far side of the COVID-19 epidemic curve should consider local strategies for re-opening and re-closing.

## 1 Introduction

Outbreak containment through testing, case isolation, contact tracing, and quarantine is often the first line of defense against a novel emerging infectious disease [2, 8, 32]. However, efforts to contain SARS-CoV-2 outbreaks have failed in many jurisdictions, leading decision-makers to supplement contact tracing with effective but socio-economically costly interventions such as school and workplace closure and other means of physical distancing [3, 15].

These measures have flattened the epidemic curve: they have reduced the effective reproduction number of SARS-CoV-2 below one, meaning that each infected case is infecting less than one person on average [3]. The epidemic curve is a common way to visualize the spread of an infectious disease and has become ubiquitous during the coronavirus disease 2019 (COVID-19) pandemic. Data on cases over time lends itself naturally to analysis by compartmental epidemic models that assume a homogeneously mixing population. Such models can be a valid approximation for many applications [22,41]. However, the epidemic curve can also obscure the spatio-temporal nature of infectious diseases, as infections jump between neighbouring populations [33]. In the early stages of an outbreak, cases are few and thus subject to random effects (stochasticity). And in the late stages of an outbreak, cases are both stochastic and spatially dispersed across multiple population centres connected through travel [18].

In such early and late stages of an epidemic, a stochastic, spatially structured model can capture important features of disease dynamics [6, 27, 28, 30]. When cases are rare, the infection may go locally extinct due to chance events–an effect referred to as stochastic fade-out [4, 9, 33]. This has nontrivial interactions with the spatial structure of the population [46]. If cases are still high in other populations, the virus may be subsequently re-introduced from those other populations through travel [14, 33]. But if the infection has also faded out in the other populations, the virus is eradicated [16].

As cases continue to decline on the far side of the COVID-19 epidemic curve, decision-makers will make choices about how and when to lift restrictions. But they will face a very different epidemiological landscape than the middle stages of the outbreak, when infections were numerous. Complete and sudden removal of these restrictions before a sufficient proportion of the population is immune to SARS-CoV-2 could cause a resurgence of cases [45]. Hence a phased approach to open or close schools and workplaces, based on “trigger” conditions such as the number of local confirmed positive cases, might be better [15, 45].

Phased approaches might be temporal in nature, with certain types of workplaces being opened before other types, for instance. Alternatively, a spatially phased approach is also possible, with smaller and/or less densely populated areas being re-opened before larger urban centers [34]. Spatially phased approaches are based on the hypothesis that during the later stages of a pandemic, the force of infection in smaller populations could be significantly less than larger populations due to more frequent stochastic fade-out [4, 9, 33], reduced contact rates on account of lower population densities [5, 21, 24, 26], and/or reduced case importation due to fewer travel connections [13]. Indeed, Ontario’s four largest cities have 2.5 times more COVID-19 cases per capita than the rest of the province (Fig. 1a) [37]. Hence, school and workplace closure could be lifted first in those populations where they provide little marginal benefit. But under a spatially phased approach, coordination between local populations remains paramount [39], given that pathogens can spread rapidly between populations during a pandemic [13, 15]. Re-importation risk may compound when local closures are poorly coordinated, with some populations eager to lift closures and hesitant to re-enact them when needed.

**Figure 1:**
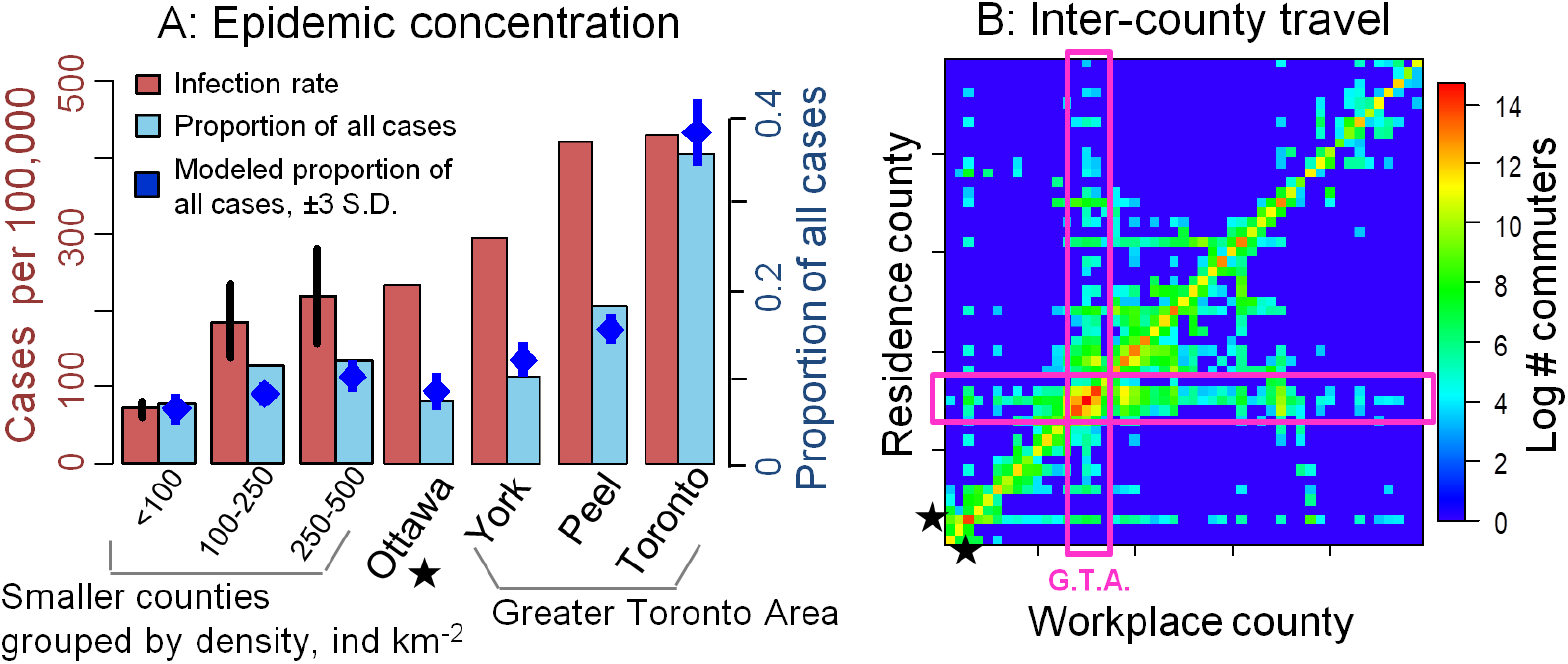
Concentration of COVID-19 cases in urban counties (a) and travel patterns among counties (b). Bars in (a) denote infection rate (cumulative cases per 100,000, red) and proportion of all cases in Ontario (blue) across counties by June 10, 2020 [37], and blue points denote the fit of 20 stochastic model realizations. Counties *<*1 million are aggregated by population density, with vertical lines denoting standard error across counties. In (b) boxes denote the Greater Toronto Area and star denotes Ottawa, data from Ref. [1].

These observations emphasize three things about modelling COVID-19 interventions on the far side of a flattened curve: (1) a stochastic transmission model might be useful for capturing disease dynamics once cases become rare; spatial structure is important for evaluating spatially phased plans, since the pathogen will not be present everywhere all the time, and (3) cases can be re-imported from other locations that have not eliminated the infection, suggesting that coordination between counties under a spatially phased approach could remain important. Our objective is to develop a spatially-structured stochastic model of SARS-CoV-2 transmission, testing, and school and workplace closure in order to address three questions: (1) Are closures best lifted at the scale of an entire province or on a county-by-county basis? (2) Does coordination of testing protocols and re-opening criteria between counties improve outcomes? (3) How well can a spatially phased approach work in the early stages of the epidemic? We use our model to determine the timing and organizational scale at which school and workplace re-opening strategies can minimize both the number of infections and person-days lost to closures, during the late-stage and early-stage epidemic. Our model is parameterized with epidemiological, demographic, and travel data for the counties, municipalities, and districts (collectively, “counties”) of Ontario, Canada.

## Results

### Model overview

We model a population distributed across local population centres (“counties”) connected through travel. Within each county, transmission follows an SEPAIR disease natural history: *S* is susceptible to infection, *E* is infected, but not yet infectious (or simply, ‘exposed’), blue*P* is pre-symptomatic infectiousness (or simply, ‘pre-symptomatic’), *A* is infectious without ever developing symptoms (or simply, ‘asymptomatic’), *I* is both infectious and symptomatic (or simply, ‘symptomatic’), and *R* is removed (no longer infectious). Symptomatic individuals are tested for SARS-CoV-2 and their status becomes ascertained with some probability per day. The infection transmission probability in a county depends on the number of contacts in schools and workplaces–which are reduced by closures–and on contacts in other settings not affected by closures, such as homes. Transmission also depends on how effectively closures reduce transmission, and the extent to which population size drives transmission. The population behavioural response to the presence of the COVID-19 is an important feature of physical distancing [11, 17, 29, 40, 47]. Hence, we assumed that transmission outside of schools and workplaces is reduced by individual physical distancing efforts (restricting social contacts, washing hands, *etc*) and that more confirmed positive cases in the county cause more individuals to practice physical distancing. Each individual travels from their home county to another county for the day with some probability (Fig. 1b) that is reduced if schools and workplaces are closed in the destination county. Additional details on model structure, data sources, parameter values, and calibration appear in Materials and Methods. Parameter definitions, values, and literature sources are summarized in SI Appendix, Table 1.

### System dynamics

We ran re-opening and re-closing simulations over a time horizon of one year and projected the number of cases in each county. Each simulation began with a 75-day period of province-wide closure applied once 325 confirmed positive cases accumulated in the province. After this period, we contrasted a “local strategy” of re-opening and re-closing counties individually according to a trigger prevalence of confirmed positive COVID-19 cases in the county, to a “global strategy” of re-opening and re-closing the entire province according to a trigger prevalence of confirmed positive COVID-19 cases in the province. Our model dynamics are characterized by two distinct regimes (Fig. 2a,c). In highly-populated counties, COVID-19 is endemic throughout the time horizon of the simulation. However, in counties with lower populations, cases blink in and out during the year, as infections jump between counties through travel and decline due to testing and voluntary distancing alone. The infection patterns appear qualitatively similar under both strategies (Fig. 2a,c), but closure patterns are very different, with most counties being closed most of the time under the global strategy (Fig. 2b,d).

**Figure 2:**
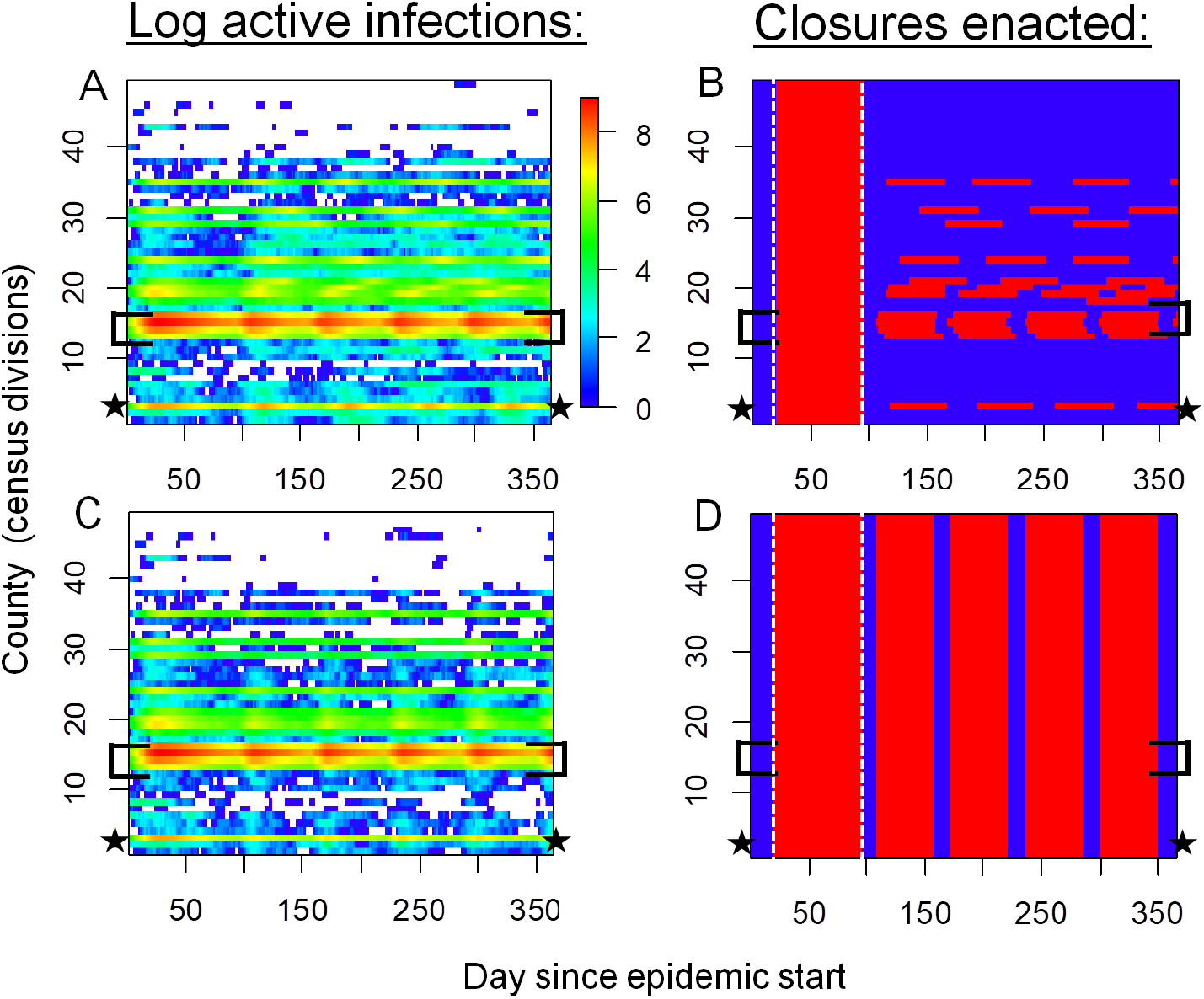
Spatiotemporal patterns of local COVID-19 cases and school/workplace closures. Panels show actual (confirmed plus not ascertained) number of COVID-19 cases (a,c) and time periods of workplace closure (red) and opening (blue) (b,d) in Ontario, Canada under local (a, b) and global (c, d) strategies. Disease dynamics are sporadic in low-population counties but endemic in high-population counties. The local strategy generates a similar disease burden to the global strategy but requires fewer person-days of closure. Optimal trigger prevalence is assumed (blue dashed lines, Fig. 3a-b). Brackets denote the Greater Toronto Area, star denotes Ottawa, and vertical dashed lines in (b, d) delineate the initial province-wide closure. All simulations were initialized with 2500 exposed persons on day 1.

### Local *versus* global re-opening strategies

The local strategy tends to outperform the global strategy for most values of the trigger prevalence (Fig. 3). When the trigger prevalence is very high (i.e. an extreme scenario where decision-makers re-open or re-close for a prevalence of 1,000 confirmed positive cases per 100,000), a high proportion of the population becomes infected, since school and workplace closures are rarely sustained in either strategy after the initial 75-day province closure. At the other extreme of the lowest trigger prevalence, both strategies minimize infections by maintaining closures for the majority of the year. However, intermediate values of the trigger prevalence represent a “sweet spot” for the local strategy, where it outperforms the global strategy in terms exhibiting significantly fewer person-days of closure for a comparable number of COVID-19 cases. The local strategy can accomplish this because it affords flexibility to enact closures only in areas with continuing active outbreaks–primarily more populous counties with higher epidemic spread rates. We identify an optimal trigger prevalence as the trigger prevalence that allows significant reductions in person-days lost to closure, but only permits cases to increase by 1% compared to its minimum value across all values of the trigger prevalence (blue dashed lines, Fig. 3). At this optimal trigger, the local strategy results in 20% fewer person-days of closure across the entire province than the global strategy.

**Figure 3:**
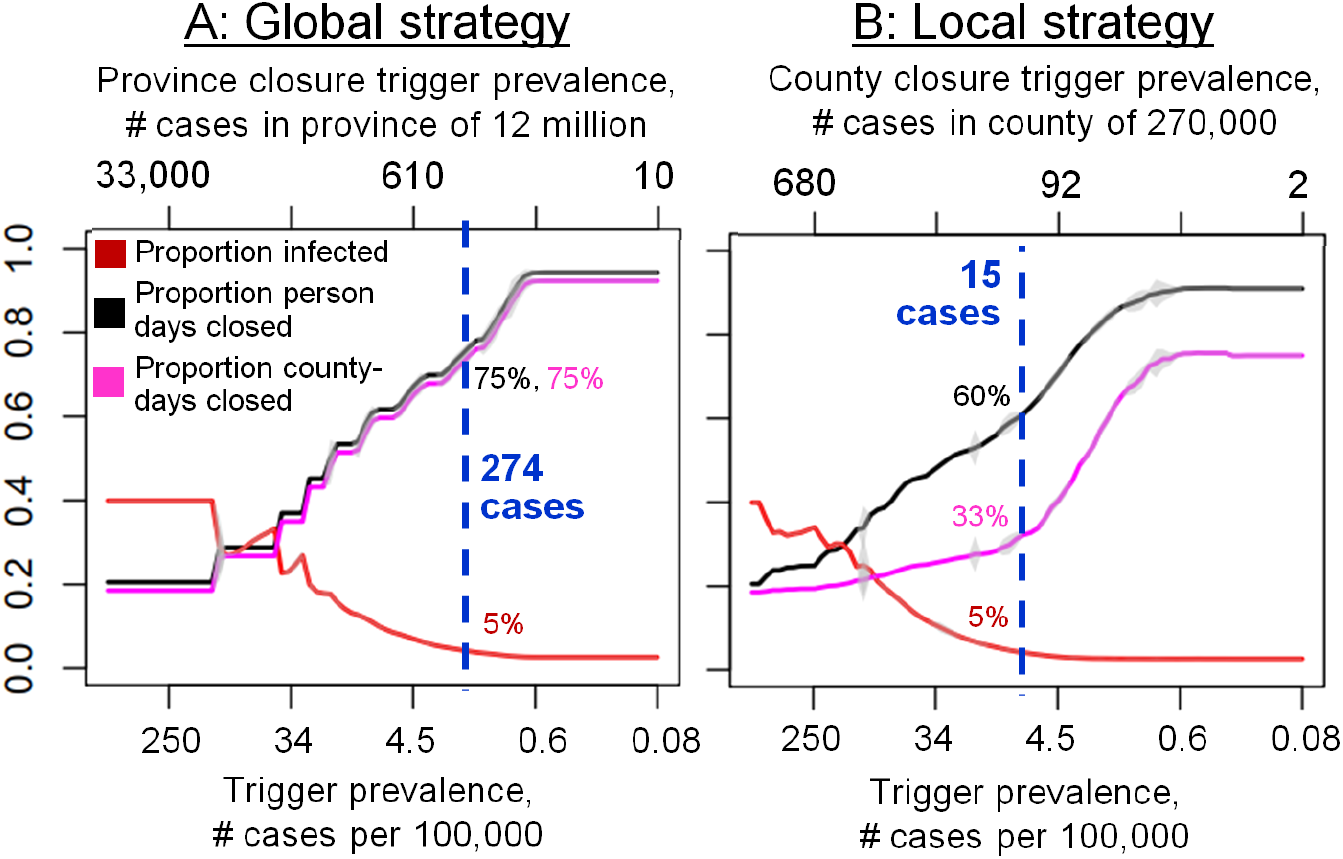
The local strategy greatly reduces person-days of school and workplace closure while only causing a small increase in the number of COVID-19 cases, relative to the global strategy. Effect of trigger prevalence on proportion infected (red) and proportion of person-days under closure (black) for the global (a) and local (b) strategy. Vertical blue lines denote optimal trigger prevalence that maintains proportion infected within 1 % of its minimum value while minimizing person days closed. Intervals represent *±*2 standard deviations across 30 model realizations.

### Benefits of coordination

A local strategy could enable different counties to adopt different triggers. Our simulation results confirm that poor coordination can undermine the benefits of the local strategy (Fig. 4a,b). As between-county variation in the trigger prevalence increases, both the mean and 85 % quantile across stochastic realizations of both the proportion infected and person-days closed rise under a broad range of assumptions for inter-county travel rates (Fig. 4a,b). The rise in infections in this scenario is somewhat counteracted by the rise in person-days lost to closure: renewed outbreaks in counties that lift closures prematurely export infections to neighbouring counties, which in turn necessitates additional closures in those counties and increases the number of person-days lost to closure (Fig. 4b). This emphasizes how close coordination can be beneficial from both public health and economic perspectives. Lack of coordination in testing is also problematic (SI Appendix, Fig. 1). As between-county variation in the testing rate for symptomatic individuals increases, the mean and 85 % quantile of proportion infected and person-days lost to closure increase in most of the stochastic realizations.

**Figure 4:**
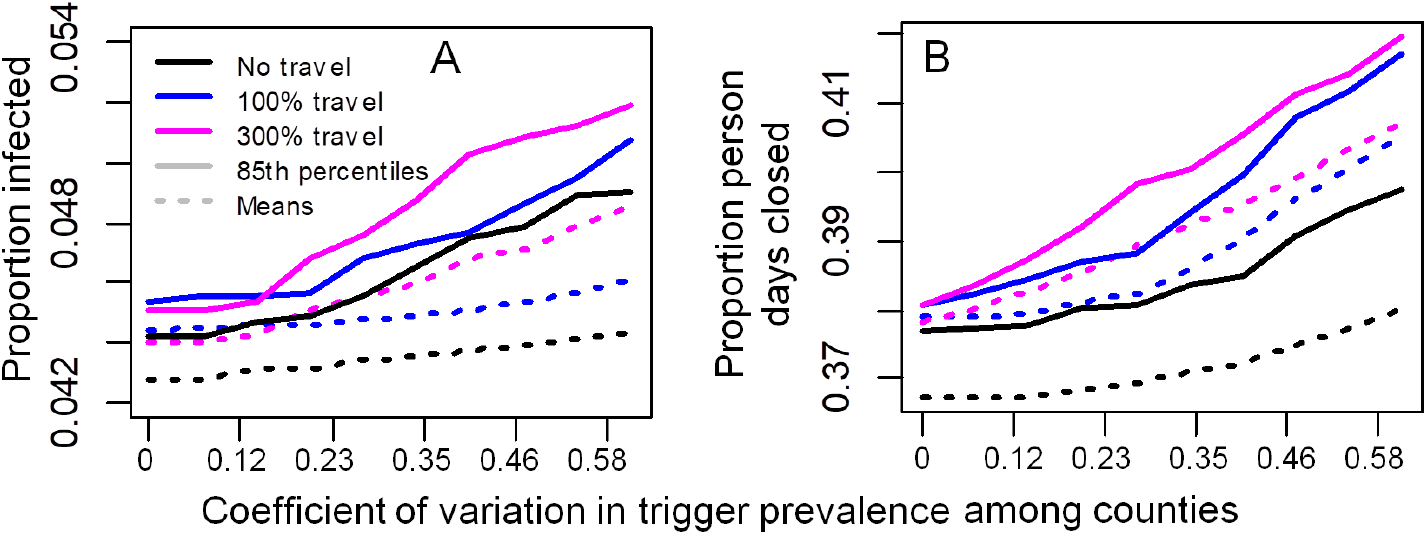
Decreasing coordination in trigger prevalence across counties increases cases and person-days closed under the local strategy. *γ*_*l,j*_ bluefollows uncorrelated variation among counties according to a uniform distribution with mean *γ*_*l*_ = 15 cases/100,000 at the optimal trigger prevalence in Fig. 3b.

### Sensitivity analysis

These results are qualitatively unchanged under moderate changes to parameter values in univariate sensitivity analyses (SI Appendix, Fig. 2). Projections are most sensitive to variation in the transmission probability, efficacy of physical distancing, and the removal rate. The performance of the local *versus* global strategies depends relatively little on the extent to which transmission probabilities are driven by population size, in other words, how rapidly the probability that a given susceptible person is infected by a given infectious person declines with the population size of the county (*ξ* is changed and model is refitted with new *ξ* values, SI Appendix, Fig. 3). Similarly, the relative performance of the two strategies is not strongly affected by doubling of travel rates (SI Appendix, Fig. 4): although cases and person-days lost to closure increase for both strategies, the local strategy retains its relative performance lead over the global strategy.

### Local closures in the early epidemic

We also compared a modified local strategy of omitting the initial 75-day provincewide closure and closing counties one at a time from the very beginning (followed by re-opening and re-closing counties as needed), to our baseline local strategy of following a 75-day province-wide closure with re-opening and re-closing counties one at a time. We found that the modified local strategy could outperform the baseline local strategy under specific conditions for trigger prevalence and testing rates (Fig. 5). In particular, the trigger prevalence must be reduced compared to our baseline analysis (Fig. 5a), such that counties are closed as soon as a few cases are detected (Fig. 5c). The optimal trigger prevalence for the modified local strategy increases exponentially with the testing probability (from 17 to 120 positive active cases in a city the size of Ottawa), meaning that counties can apply less stringent triggers only if their testing rates are very high and find more cases (Fig. 5c). Testing of asymptomatic individuals is not included in our baseline analysis, but might occur under high testing capacity and effective contact tracing, and would permit a higher trigger prevalence and reduced person-days closed under the modified local strategy (Fig. 5b) by limiting epidemic growth to the most populous counties. However, given the initial short supply of test kits and long testing turnaround times that characterized Ontario and many other jurisdictions, the testing rate for symptomatic individuals probably remained below the required 0.1/day during the early epidemic in Ontario. Sensitivity analysis in the early epidemic (Fig. 5b) additionally shows that the benefit of fewer person-days closed under the local strategy declines when closures begin after many thousands of people are already infected, when travel is high, or if initial infections are concentrated in cities. Taken together, these results suggest that the modified local strategy of omitting the 75-day province-wide closure could significantly outperform the baseline local strategy early in the initial epidemic only with prompt mitigation, moderate-to-high testing rates, and very low trigger prevalence (a scenario resembling the South Korean control strategy). This finding reiterates public health consensus that early and aggressive action in the early stages of a pandemic, and also potentially during second waves, could minimize both infections and total person-days of closure.

**Figure 5:**
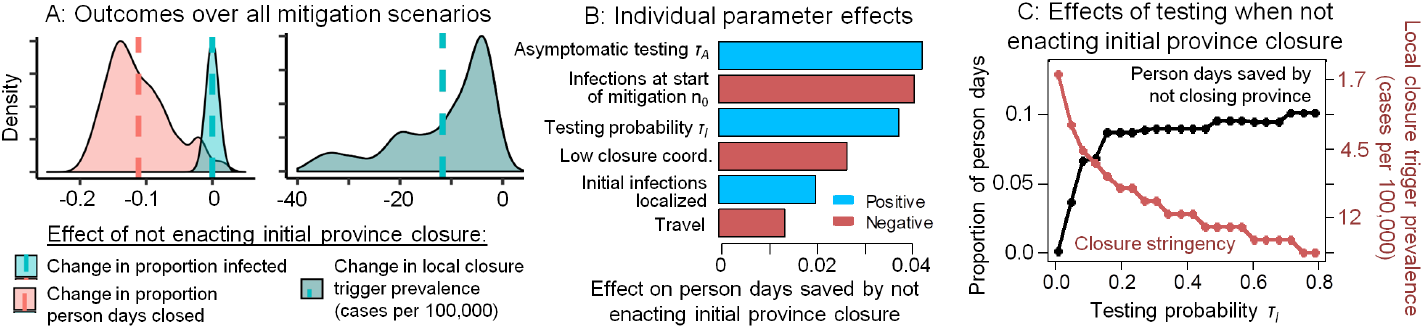
Using county-by-county closures from the beginning and omitting the initial 75-day province-wide closure at the start of an epidemic can minimize infections and person-days of closure under moderate-to-high testing rates and low trigger prevalence. (A) Across all combinations of control parameters (specified in B), not enacting an initial 75-day province closure at the start of the epidemic reduces person-days closed but requires a lower trigger prevalence in county-level closure decisions (vertical lines denote means). (B) Sensitivity analysis of how each control parameter affects person-days of closure avoided by omitting the initial 75-day provincewide closure. (C) Moderate-high testing levels allow less stringent county closure criteria (trigger prevalence, red) and results in fewer person-days closed (black) compared to 75-day province-wide closure. Person-days lost, infections, and trigger prevalence calculated over the first 120 days of the epidemic. Throughout this analysis, we use a baseline 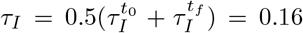 In (A) and (B) *τ*_*I*_ and _0_ were varied over *±*75% of their baseline values, low coordination is either absent or present (in which case *γ*_*l,j*_ among counties follows a uniform distribution with coefficient of variation 0.27), asymptomatic testing *τ*_*A*_ = 0 or *τ*_*A*_ = *τ*_*I*_*/*2 = 0.08, and initial infections are either distributed evenly among the population or concentrated in two randomly selected counties population *>* 500, 000.

## Discussion

Plans for re-opening and re-closing schools and workplaces in the later stages of COVID-19 epidemics are diverse and uncoordinated. Some re-opening guidelines include epidemiological triggers such as case incidence or contact tracing capacity [36], while others include guidelines for re-opening on a county-by-county basis [34]. Our results suggest that plans for re-opening economies on the far side of the COVID-19 epidemic curve should consider preceding larger-scale re-openings with local re-openings. However, for this to work, the trigger conditions need to be coordinated by the province: individual counties cannot draw up guidelines independently.

Our model was parameterized for the province of Ontario, Canada. Sensitivity analysis showed our results were robust to assumptions regarding transmission processes and travel patterns. The robustness of these results stems from being able to re-open more sparsely populated counties that have a lower case burden and can benefit from stochastic fade-out more often than densely-populated urban centres (Fig. 2). In turn, this suggests that the results may apply more broadly to other Canadian provinces and US states with similarly low population density and dispersed spatial structure. However, this would need to be confirmed with model extensions that are tailored to these other jurisdictions. Additionally, not all US states began their control efforts with a period of closure that was effective enough to flatten their epidemic curves, which is a crucial difference.

Province- and state-wide lockdowns have generated resistance from populations that feel the restrictions should not apply to them. This outcome is tied up with the phenomenon of policy resistance, where the nonlinear behavioural response to an intervention partly undermines the intervention [43]. Nonlinear interaction between human and natural systems is pervasive [25], and epidemiological systems are no exception [7, 47]. Behavioural feedback during the COVID-19 pandemic has manifested through (1) physical distancing to minimize individual infection risk in response to rising case reports [40], and (2) pushback against lockdowns on account of economic impacts or perceived restrictions on civil liberties. An evidence-informed and coordinated approach to lifting lockdowns in less densely populated areas first, such as the one we propose, might have the added benefit of improving compliance to measures in populations that perceive province-wide closure to cover a needlessly large section of the population. In a related vein, local closures may be more effective if local decision-makers can enact closures more promptly than what is possible under a province-wide decision-making process.

Our model did not include several features that could influence predictions. A key assumption was that individual counties enact closures as soon as positive cases exceed a trigger prevalence. In practice, a delay could allow case notifications to surge. Hesitation to re-close counties as needed can be especially problematic because (1) an increase in deaths follows weeks behind an increase in cases, and (2) the optimal trigger prevalence found here translates to closing counties when only a few active cases are detected (Fig. 3b). Future iterations of this model could include other important features such as age structure [45] or ICU capacity. These features could enable projecting the effect of re-opening only primary and elementary schools, or using local ICU occupancy as a trigger. Future work could also explore the role of extra-provincial case importations in the late stage of the pandemic, which could become important once local cases become rare [33].

Data on SARS-CoV-2 epidemiology, interventions and treatments will become both increasingly available and increasingly reliable as the COVID-19 pandemic unfolds. There is a corresponding urgent need to develop more detailed models that can address a broader range of policy questions, so that evidence-based policy-making has more information upon which to base decisions. Stochastic, spatially structured models may become increasingly useful for informing re-opening and re-closing strategies in the COVID-19 pandemic, by allowing decision-makers to explore the potential advantages of coordinated county-by-county strategies.

## Methods

The following subsections describe the details of the model structure and parameterization. A table of parameter definitions, baseline values, and literature sources appears in SI Appendix: Table 1.

### Population structure and travel

The model contains 49 local populations (“counties”) that represent each of 49 Ontario census divisions and have the same corresponding population sizes [1]. At the start of each day, each individual in county *j* travels to county *k* for the day with probability *m*_*jk*_, in which case they experience any transmission events that occur there. At the end of the day, they return back to county *j*. The values of the travel matrix **M** = [*m*_*jk*_] were obtained from survey data [1] spanning 39% of Ontario inhabitants, of whom 25% worked outside their census division, arriving at an aggregate daily travel probability of 10% per day. This likely overestimates the impact of travel since we are treating each individual in the population as equally likely of travelling to another county each day, whereas in real populations, the same individuals tend to travel each day and tend to repeat their contacts with the same individuals. Infected individuals are less likely to travel by a factor *r* = 0.19 (i.e. 19% less likely to travel), since 81% of reported COVID-19 cases are mild [12]. Individuals who test positive are less likely to travel by a factor *η* = 0.8 [20, 42], with travel by individuals sick and confirmed positive reduced by (1 − *η*)(1 − *r*). Additionally, each individual’s travel probability to a closed county is reduced by a factor *E*, since fewer individuals travel to a county when its schools and workplaces are closed.

### Transmission and testing

The state *{D*_*i*_, *T*_*i*_*}* of individual *i* reflects both their epidemiological status *D*_*i*_ *∈ {S, E, A, P, I, R}* and their testing status, where *T*_*i*_ *∈ {N, K}* for not known/known infection status, respectively. *{·, T*_*i*_*}* denotes an individual with testing status *T*_*i*_ and any of the five epidemiological states, with similar interpretation for {*D*_*i*_, *·*}. *P*_*j*_ denotes the population size of county *j* and 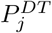 denotes the number of individuals of state {*D, T}* in county *j*. Each timestep lasts one day. During each day, each individual’s epidemiological status in county *j* is updated as follows:

1. Individuals in the *{S, ·}* state become exposed with probability *λ*_*j*_, entering the *{E, ·}* state.
2. blueIndividuals in the *{E, ·}* state become pre-symptomatic with probability (1 − *π*)*α* and enter the *{P, ·}* state, or become asymptomatic with probability *πα* and enter the *{A, ·}* state.
3. Individuals in the *{P, ·}* state become symptomatic with probability *σ*, entering the *{I, ·}* state.
4. Individuals in the *{I, N}* state are tested with probability *τ*_*I,j*_, entering the *{I, K}* states.
5. blueIndividuals in the *{I, ·}* and *{A, ·}* states are removed (are no longer infectious) with probability *ρ*, entering the *{R, ·}* state.

Infection history parameter values are obtained from epidemiological literature [35, 44]. We assign each newly infected individual to be a super-spreader with probability *s* = 0.2 [31] and denote super-spreading (non-super-spreading) individuals with the subscript *s* (e.g., *A*_*s*_, *I*_*s*_) (*ns*, resp.). Super-spreaders infect others with a probability that is (1 − *s*)*/s* times higher than non-superspreaders. blueDaily testing probabilities improved over the the first 60 days of province-wide closures as processing time declined three-fold and testing increased 100-fold [38]. To model this, we assumed daily testing probabilities increase from an initial level 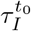 to 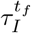 by day 60 after the 325th positive case is detected and constrained 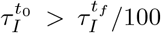. Physical distancing through closures or behavioural changes can reduce the probability of transmission by up preventing a fraction of all contacts. Closure *C*_*j*_(*t*) (see below) of schools and workplaces in county *j* can be applied and lifted over time and affect a fraction *w* of all contacts. We take *w* = 0.45 based on time use data for time spent at schools, workplaces, and other institutions that can be mandated to close [10]. The remaining time spent, 1 − *w*, is in settings such as homes and social gatherings. We assumed that individuals reduce their contacts in proportion to the number of confirmed cases reported in their county by a factor 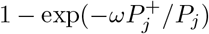, where 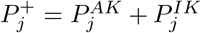 Hence, the fraction of contacts *F*_*j*_(*t*) remaining after physical distancing in county *j* at time *t* is therefore

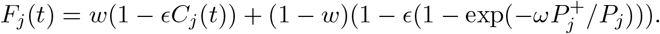

The contacts of an infected person decline from *f*_*T* =*N*_ = 1 to a fraction *f*_*T* =*K*_ = 1-*η* if they test positive for COVID-19, where *η* = 0.8 [20, 42]. The transmission probability also depends on the individual’s epidemiological state *D*, with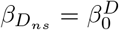 for non-superspreaders *{A*_*ns*_, *·}, {I*_*ns*_, *·}* and 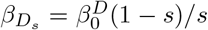 for superspreaders {*A*_*s*_}, {*I*_*s*_,.}. Hence the probability per day that a susceptible person in county *j* is infected by an infectious person is:

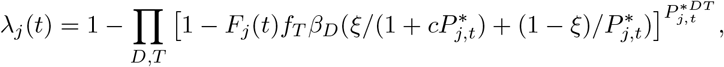

since this is 1 minus the probability of not being infected by any class of the infectious individuals in county *j*. The starred notation, 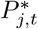 (resp. 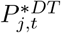) denotes the population size (resp. number of individuals of disease status *D* and testing status *T*) on day *t* after adjusting for travel. blue*ξ* and *c* control how the transmission probability depends on population size: standard incidence is recovered for *ξ* = 0, while *ξ* = 1 recovers a scenario where contacts increase with population size to an extent controlled by *c* [5].

### School and workplace closure

In Ontario, an emergency was declared on the day the cumulative number of reported positive cases *t*_*n*_ reached 325 (March 17), leading to closures of workplaces (schools were already closed for March Break). Hence, closure strategies in our model were enacted only after an initial *t*_*start*_ = 75-day province-wide closure expires. Re-openings (and re-closures) are subsequently enacted under the local and global strategy when the percentage of confirmed cases fall below (or exceed), a trigger prevalence *γ*_*G*_ at the province level or *γ*_*l,j*_ within a county *j*. Under limited coordination, *γ*_*l,j*_ may be greater in counties eager to lift or hesitant to enact closures. Any closures last for *δ*_*C*_ = 50 days, with *t*_*G*_ (*t*_*l,j*_) denoting the last time a closure was enacted in the province (in county *j*), after which period the closure decision is re-evaluated. The closure function *C*_*j*_(*t*) is then

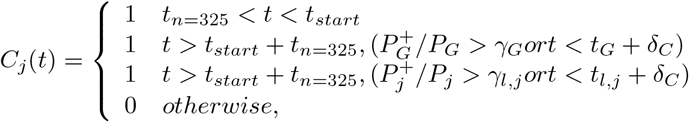

where 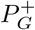 is the total number of known, active cases in the province and *P*_*G*_ is province population size.

### Calibration

We set 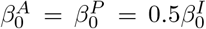 [19], based on data showing 44% of SARS-CoV-2 shedding occurs before symptoms develop and our assumed duration of infectious periods. blueGiven that contact rates can depend on population size, we determined 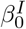 under each value of *ξ* and *c* by calibrating the model in the absence of distancing, closure, and testing such that 65% of the population is infected, based on the assumption that *R*_0_ = 2.3 and using resulting projections of the final size from compartmental epidemic models [23, 45]. We calibrated intervention efficacy (, *ω*), testing 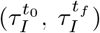, and transmission dependence on population size (*ξ, c*) by fitting model outputs of (1) timeseries of incident confirmed cases (the number of individuals entering the *I, K* state each day) to empirical data on daily confirmed cases by reporting date [37] (SI Appendix, Fig. 5); (2) the spatial distribution of cumulative confirmed positive cases by day 85 of the outbreak (June 10, 2020) in the model to data [37]; (3) the modelled ratio of actual cases to confirmed positive cases province-wide (i.e. number of individuals not in *S*, to the cumulative number of individuals tested positive), to an empirical estimate of this ratio of 8.76 for under-ascertainment in the US lachmann2020correcting (8.7 *±* 2.3 in our model, mean*±*2 S.D.); and (4) the modelled amount of discretionary physical distancing 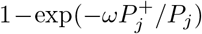 by day *t* − *t*_*n*=325_ = 21 in our simulation to an empirical estimate of ≈ 50% adherence to physical distancing by day 21 of the outbreak (April 6, 2020) from a public survey gallup1 (0.49 ± 0.3 in our model). blueIn fitting the spatial case distribution, we calculated the proportion of all confirmed positive tests found in the 4 most populous Ontario cities and in 3 sets of counties grouped by population density. We adjusted the proportion of total cases and total cases per 100,000 (Fig. 1a) observed in each county or county group *j* for spatial differences in the number of tests *T*_*j*_ per capita, by multiplying the observed number of cases in each location by the fraction (*P*_*j*_ ∑ *T*_*j*_)/ (*T*_*j*_ ∑ *P*_*j*_) with fraction values ranging 0.72-1.16).

This research was supported by the Natural Sciences and Engineering Research Council (MA, CTB).

## Data Availability

The model simulation code is available upon request. All other data used in the model are publicly available.

## Notes

### Competing Interest Statement

The authors have declared no competing interest.

### Funding Statement

This research was funded by NSERC Discovery Grants to CTB and MA. The funder had no role in the research.

## References

[1] Statistics canada, 2016 census, catalogue no. 98-400-x2016391., Accessed April 7, 2020.

[2] Alberto Aleta, David Martin-Corral, Ana Pastore y Piontti, Marco Ajelli, Maria Litvinova, Matteo Chinazzi, Natalie E Dean, M Elizabeth Halloran, Ira M Longini Jr, Stefano Merler, et al. Modeling the impact of social distancing, testing, contact tracing and household quarantine on second-wave scenarios of the covid-19 epidemic. medRxiv, 2020.

[3] Sean C Anderson, Andrew M Edwards, Madi Yerlanov, Nicola Mulberry, Jessica Stockdale, Sarafa A Iyaniwura, Rebeca C Falcao, Michael C Otterstatter, Michael A Irvine, Naveed Z Janjua, et al. Estimating the impact of covid-19 control measures using a bayesian model of physical distancing. medRxiv, 2020.

[4] Håkan Andersson and Tom Britton. Stochastic epidemics in dynamic populations: quasi-stationarity and extinction. Journal of mathematical biology, 41(6):559–580, 2000.

[5] Janis Antonovics, Yoh Iwasa, and Michael P Hassell. A generalized model of parasitoid, venereal, and vector-based transmission processes. The American Naturalist, 145(5):661–675, 1995.

[6] Julien Arino, Jonathan R Davis, David Hartley, Richard Jordan, Joy M Miller, and P Van Den Driessche. A multi-species epidemic model with spatial dynamics. Mathematical Medicine and Biology, 22(2):129–142, 2005.

[7] Chris T Bauch and Alison P Galvani. Social factors in epidemiology. Science, 342(6154):47–49, 2013.

[8] Chris T Bauch, James O Lloyd-Smith, Megan P Coffee, and Alison P Galvani. Dynamically modeling sars and other newly emerging respiratory illnesses: past, present, and future. Epidemiology, pages 791–801, 2005.

[9] Michael Boots and Akira Sasaki. Parasite evolution and extinctions. Ecology Letters, 6(3):176–182, 2003.

[10] Bureau of Labor Statistics. American time use survey — 2018 results. https://www.bls.gov/news.release/pdf/atus.pdf, 2018. accessed 4 April 2020.

[11] Vincenzo Capasso and Gabriella Serio. A generalization of the kermack-mckendrick deterministic epidemic model. Mathematical Biosciences, 42(1-2):43–61, 1978.

[12] CDC. Severe outcomes among patients with coronavirus disease 2019 (COVID-19)—united states, february 12–march 16, 2020. 2020.

[13] Vittoria Colizza, Alain Barrat, Marc Barthelemy, Alain-Jacques Valleron, and Alessandro Vespignani. Modeling the worldwide spread of pandemic influenza: baseline case and containment interventions. PLoS Med., 4(1), 2007.

[14] Gaston De Serres, Nigel J Gay, and C Paddy Farrington. Epidemiology of transmissible diseases after elimination. American Journal of Epidemiology, 151(11):1039–1048, 2000.

[15] Sara Y Del Valle, Susan M Mniszewski, and James M Hyman. Modeling the impact of behavior changes on the spread of pandemic influenza. In Modeling the interplay between human behavior and the spread of infectious diseases, pages 59–77. Springer, 2013.

[16] David JD Earn, Pejman Rohani, and Bryan T Grenfell. Persistence, chaos and synchrony in ecology and epidemiology. Proceedings of the Royal Society of London. Series B: Biological Sciences, 265(1390):7–10, 1998.

[17] Eli P Fenichel, Carlos Castillo-Chavez, M Graziano Ceddia, Gerardo Chowell, Paula A Gonzalez Parra, Graham J Hickling, Garth Holloway, Richard Horan, Benjamin Morin, Charles Perrings, et al. Adaptive human behavior in epidemiological models. Proceedings of the National Academy of Sciences, 108(15):6306–6311, 2011.

[18] Neil M Ferguson, Christl A Donnelly, and Roy M Anderson. Transmission intensity and impact of control policies on the foot and mouth epidemic in great britain. Nature, 413(6855):542–8, 2001.

[19] Xi He, Eric HY Lau, Peng Wu, Xilong Deng, Jian Wang, Xinxin Hao, Yiu Chung Lau, Jessica Y Wong, Yujuan Guan, Xinghua Tan, et al. Temporal dynamics in viral shedding and transmissibility of COVID-19. medRxiv, 2020.

[20] Joel Hellewell, Sam Abbott, Amy Gimma, Nikos I Bosse, Christopher I Jarvis, Timothy W Russell, James D Munday, Adam J Kucharski, W John Edmunds, Fiona Sun, et al. Feasibility of controlling covid-19 outbreaks by isolation of cases and contacts. The Lancet Global Health, 2020.

[21] Samuel Heroy. Metropolitan-scale COVID-19 outbreaks: how similar are they? arXiv preprint 2004.01248, 2020.

[22] Herbert W Hethcote. The mathematics of infectious diseases. SIAM review, 42(4):599–653, 2000.

[23] Joe Hilton and Matt J Keeling. Estimation of country-level basic reproductive ratios for novel coronavirus (covid-19) using synthetic contact matrices. medRxiv, 2020.

[24] Hao Hu, Karima Nigmatulina, and Philip Eckhoff. The scaling of contact rates with population density for the infectious disease models. Mathematical biosciences, 244(2):125–134, 2013.

[25] Clinton Innes, Madhur Anand, and Chris T Bauch. The impact of human-environment interactions on the stability of forest-grassland mosaic ecosystems. Scientific reports, 3(1):1–10, 2013.

[26] MCD Jong, O Diekmann, and H Heesterbeek. How does transmission of infection depend on population size? epidemic models. Publication of the Newton Institute, pages 84–94, 1995.

[27] Matthew J Keeling and Bryan T Grenfell. Disease extinction and community size: modeling the persistence of measles. Science, 275(5296):65–67, 1997.

[28] Grace PS Kwong and Rob Deardon. Linearized forms of individual-level models for large-scale spatial infectious disease systems. Bulletin of Mathematical Biology, 74(8):1912–1937, 2012.

[29] Alexander Lachmann. Correcting under-reported covid-19 case numbers. medRxiv, 2020.

[30] Alun L Lloyd and Robert M May. Spatial heterogeneity in epidemic models. J. Theor. Biol., 179(1):1–11, 1996.

[31] James O Lloyd-Smith, Sebastian J Schreiber, P Ekkehard Kopp, and Wayne M Getz. Superspreading and the effect of individual variation on disease emergence. Nature, 438(7066):355–359, 2005.

[32] Ira M Longini, Azhar Nizam, Shufu Xu, Kumnuan Ungchusak, Wanna Han- shaoworakul, Derek AT Cummings, and M Elizabeth Halloran. Containing pandemic influenza at the source. Science, 309(5737):1083–87, 2005.

[33] C Jessica E Metcalf, Katie Hampson, Andrew J Tatem, Bryan T Grenfell, and Ottar N Bjørnstad. Persistence in epidemic metapopulations: quantifying the rescue effects for measles, mumps, rubella and whooping cough. PloS one, 8(9), 2013.

[34] Lawrence Mower and Mary Ellen Klas. Governor announces reopening plan for state, but south florida isn’t included yet. https://www.miamiherald.com/news/politics-government/state-politics/article242368531.html, 2020; accessed April 30, 2020.

[35] Hiroshi Nishiura, Natalie M Linton, and Andrei R Akhmetzhanov. Serial interval of novel coronavirus (2019-ncov) infections. medRxiv, 2020.

[36] Ontario. A framework for re-opening our province. https://files.ontario.ca/mof-framework-for-reopening-our-province-en-2020-04-27.pdf, 2020; accessed April 30, 2020.

[37] Ontario. Ontario covid-19 data tool https://www.publichealthontario.ca/en/data-and-analysis/infectious-disease/covid-19-data-surveillance/covid-19-data-tool, 2020; accessed June 10, 2020.

[38] Ontario. The 2019 novel coronavirus (covid-19). https://www.ontario.ca/page/2019-novel-coronavirussection-0, 2020; accessed March 31, 2020.

[39] World Health Organization et al. Who global influenza preparedness plan: the role of who and recommendations for national measures before and during pandemics. Technical report, World Health Organization, 2005.

[40] Timothy C Reluga. Game theory of social distancing in response to an epidemic. PLoS computational biology, 6(5), 2010.

[41] Dieter Schenzle. An age-structured model of pre-and post-vaccination measles transmission. Mathematical Medicine and Biology: A Journal of the IMA, 1(2):169–191, 1984.

[42] FA Soud, MM Cortese, AT Curns, PJ Edelson, RH Bitsko, HT Jordan, AS Huang, JM Villalon-Gomez, and GH Dayan. Isolation compliance among university students during a mumps outbreak, kansas 2006. Epidemiology & Infection, 137(1):30–37, 2009.

[43] John D Sterman. Learning from evidence in a complex world. Am. J. Pub. Health, 96(3):505–14, 2006.

[44] Lauren Tindale, Michelle Coombe, Jessica E Stockdale, Emma Garlock, Wing Yin Venus Lau, Manu Saraswat, Yen-Hsiang Brian Lee, Louxin Zhang, Dongxuan Chen, Jacco Wallinga, et al. Transmission interval estimates suggest pre-symptomatic spread of COVID-19. medRxiv, 2020.

[45] Ashleigh R Tuite, David N Fisman, and Amy L Greer. Mathematical modelling of covid-19 transmission and mitigation strategies in the population of ontario, canada. CMAJ, 2020.

[46] Cåcile Viboud, Ottar N Bjørnstad, David L Smith, Lone Simonsen, Mark A Miller, and Bryan T Grenfell. Synchrony, waves, and spatial hierarchies in the spread of influenza. science, 312(5772):447–451, 2006.

[47] Zhen Wang, Michael A Andrews, Zhi-Xi Wu, Lin Wang, and Chris T Bauch. Coupled disease–behavior dynamics on complex networks: A review. Physics of life reviews, 15:1–29, 2015.

